# Sino-US-DrugQA: A Benchmark for Evaluating Large Language Models in Cross-Jurisdictional Pharmaceutical Regulation

**DOI:** 10.64898/2026.02.13.26346236

**Authors:** Xuejing Fu, Zhen Chen, Wentao Lu

**Affiliations:** Information Application Research Center of Shanghai Municipal Administration for Market Regulation, Shanghai, China; Shanghai Artificial Intelligence Research Institute Co., Ltd., Shanghai, China

## Abstract

Cross-jurisdictional pharmaceutical compliance requires comparative analysis of regulatory requirements across jurisdictions such as the US FDA and China’s NMPA. Although large language models (LLMs) are increasingly explored for healthcare-related applications, their performance in cross-jurisdictional regulatory comparison has not been systematically characterized using dedicated benchmarks. This study introduces Sino-US-DrugQA, a bilingual benchmark dataset designed to evaluate LLM performance in cross-jurisdictional pharmaceutical regulation. We constructed a bilingual corpus of 11,871 multiple-choice question–answer pairs derived from authoritative NMPA regulations and US CFR Title 21. The dataset includes monolingual retrieval tasks and cross-jurisdictional comparative reasoning tasks. We conducted baseline evaluations of four representative LLMs (GPT-5.2, Gemini-3-flash, Qwen-3-235B, DeepSeek-V3.2) under a standardized zero-shot protocol with temperature set to 0. The dataset and evaluation scripts are released as open resources to support reproducible benchmarking. Across 11,871 questions, the evaluated models achieved overall accuracies ranging from 78.97% to 84.51% in the zero-shot setting. Across models, performance consistently decreased on comparative questions relative to monolingual questions (approximately 6–9 percentage points), a gap that persisted even for the strongest-performing system, highlighting cross-jurisdictional alignment and comparison-specific deduction as key challenges. Sino-US-DrugQA provides a practical resource for benchmarking regulatory AI in a high-stakes compliance setting. Current LLMs show utility as drafting and screening assistants for monolingual regulatory queries, but their limitations in cross-jurisdictional comparative reasoning support a conservative deployment posture requiring expert review. The dataset and evaluation scripts are available at https://github.com/DodgeLU/Sino-US-DrugQA.

## 1. Introduction

The globalization of pharmaceutical research and development has necessitated harmonization of regulatory standards across major jurisdictions [1,2]. Despite efforts by organizations such as the International Council for Harmonisation of Technical Requirements for Pharmaceuticals for Human Use (ICH), significant disparities persist between the regulatory frameworks of the United States Food and Drug Administration (FDA) and China’s National Medical Products Administration (NMPA) [3,4]. For multinational pharmaceutical enterprises, simultaneous submission of new drug applications in both jurisdictions - commonly referred to as “dual filing” - requires systematic, provision-level comparative analysis of Title 21 of the Code of Federal Regulations (CFR) and Chinese drug administration laws. This process extends beyond literal translation and demands an understanding of regulatory intent, procedural timelines, and jurisdiction-specific technical requirements that may differ substantially between jurisdictions. Such comparative analysis requires not only regulatory literacy but also fine-grained reasoning over non-equivalent administrative rules across jurisdictions.

Currently, cross-jurisdictional compliance relies heavily on manual gap analyses performed by regulatory affairs (RA) professionals [5,6]. This approach is labor-intensive, costly, and prone to human error, particularly given the rapid evolution of regulatory landscapes, especially in China [7]. The emergence of artificial intelligence (AI) technologies, particularly large language models (LLMs), has shown promise in supporting clinical decision-making and regulatory tasks [8,9]. LLMs can process vast amounts of multilingual text and perform complex reasoning, suggesting potential utility in regulatory technology applications such as regulatory intelligence, drafting assistance, and preliminary compliance screening [10]. However, whether such capabilities extend reliably to cross-jurisdictional regulatory comparison remains an open question.

However, deploying LLMs in cross-jurisdictional regulatory compliance presents unique challenges that distinguish it from general medical or legal question answering. Unlike clinical diagnosis or single-jurisdiction legal interpretation, cross-jurisdictional regulatory reasoning requires models to operate across partially overlapping but non-isomorphic administrative systems. A model must not only translate language but also perform concept-level alignment across regulatory systems (e.g., mapping “IND” in the US to “Clinical Trial Approval” in China) and reason over jurisdictional divergences in technical and procedural requirements, as well as differences in how regulatory obligations are specified or operationalized (e.g., role-specific personnel qualifications or mandatory written procedures). Errors in this setting carry direct compliance implications, potentially leading to application rejections, delayed market entry, or legal penalties [11]. Consequently, the reliability of LLMs in this specific cross-jurisdictional regulatory setting remains a critical, yet insufficiently validated, variable.

The application of LLMs in healthcare regulation is currently hindered by a lack of specialized benchmarks that reflect this complexity. Existing legal NLP benchmarks, such as LawBench [12] or LexGLUE [13], primarily evaluate monolingual statutory interpretation, case law reasoning, or judgment prediction within a single legal system. Similarly, medical benchmarks such as MedMCQA [14] emphasize clinical knowledge rather than administrative regulation. As a result, these benchmarks are not designed to evaluate cross-jurisdictional administrative reasoning, where models must explicitly compare non-equivalent regulatory requirements across distinct legal frameworks. This includes tasks such as identifying differences in procedural obligations, compliance thresholds, or retention requirements between jurisdictions (e.g., “Is the document retention period for clinical trials longer in the US or China?”) [15].

This gap is significant because strong performance within a single jurisdiction does not necessarily translate to reliable cross-jurisdictional regulatory reasoning, and models may generate misleading or incorrect outputs when tasked with comparative regulatory analysis [8,9]. To address this gap, we introduce Sino-US-DrugQA, a bilingual benchmark dataset specifically designed to evaluate LLM performance in cross-jurisdictional pharmaceutical regulation between China and the United States. Distinct from existing legal or medical benchmarks, Sino-US-DrugQA explicitly targets comparative administrative reasoning, rather than single-jurisdiction understanding. Rather than proposing a new model or optimization strategy, this work provides a standardized evaluation framework and a publicly available benchmarking resource for the regulatory science community. By benchmarking multiple state-of-the-art LLMs, we quantify the performance gap between monolingual regulatory understanding and cross-jurisdictional comparative reasoning, thereby offering empirical evidence for the safe, expert-supervised deployment of LLMs in global pharmaceutical regulatory practice [16].

**Table 1:**
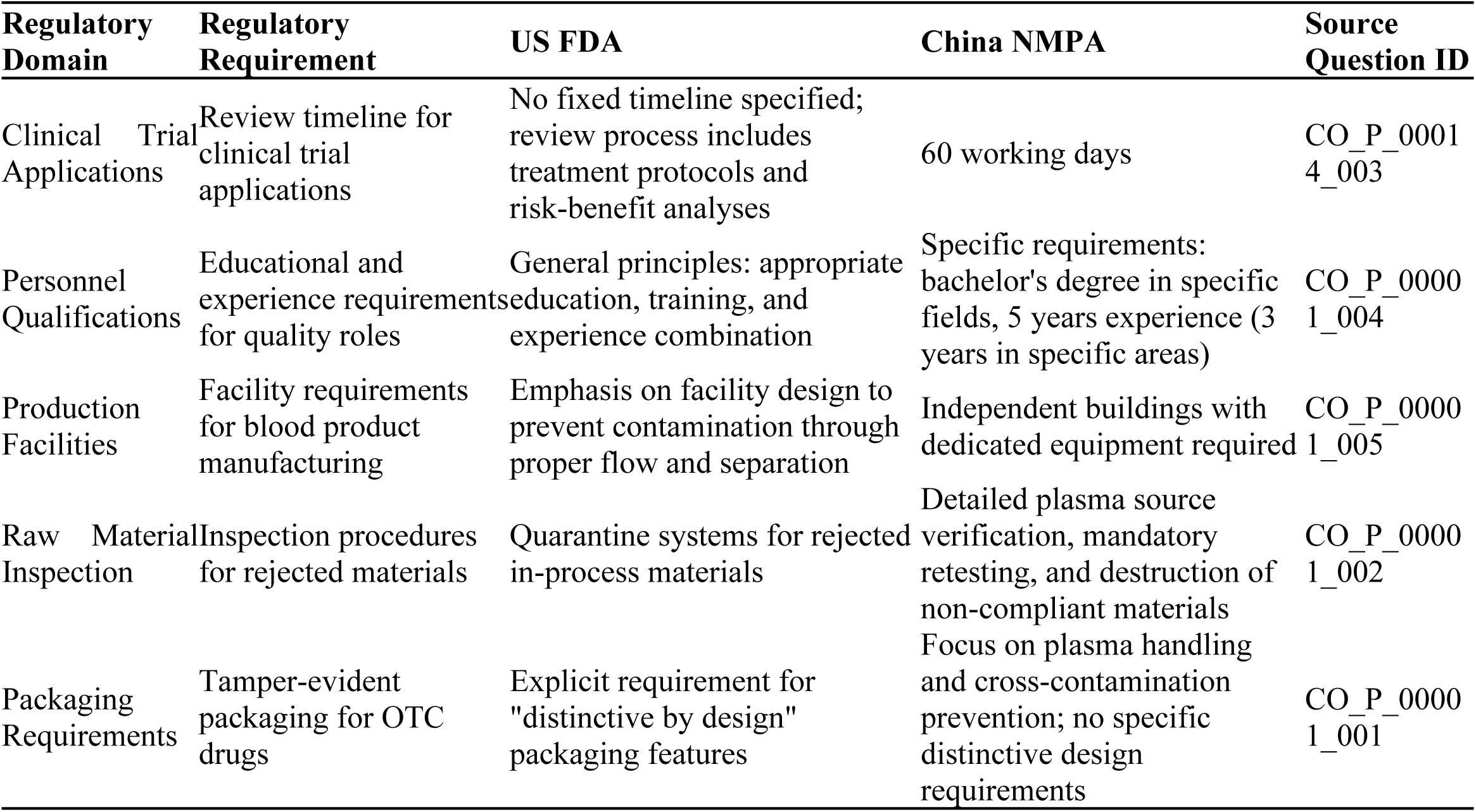
Real-world examples of regulatory divergence.

## 2. Materials and Methods

### 2.1. Regulatory Data Sources

This retrospective benchmarking study was constructed using authoritative regulatory documents issued by the NMPA and the US FDA, with document versions valid as of December 2025. To ensure comprehensive coverage of regulatory requirements, we selected documents spanning the full pharmaceutical product lifecycle, from clinical trial authorization to post-market surveillance.

The corpus comprised 134 key regulations from the NMPA and 195 documents from Title 21 of the US Code of Federal Regulations (CFR). Table 2 summarizes the primary regulatory sources used in dataset construction, illustrating functional correspondence across regulatory domains rather than asserting one-to-one legal equivalence between the two jurisdictions.

**Table 2:**
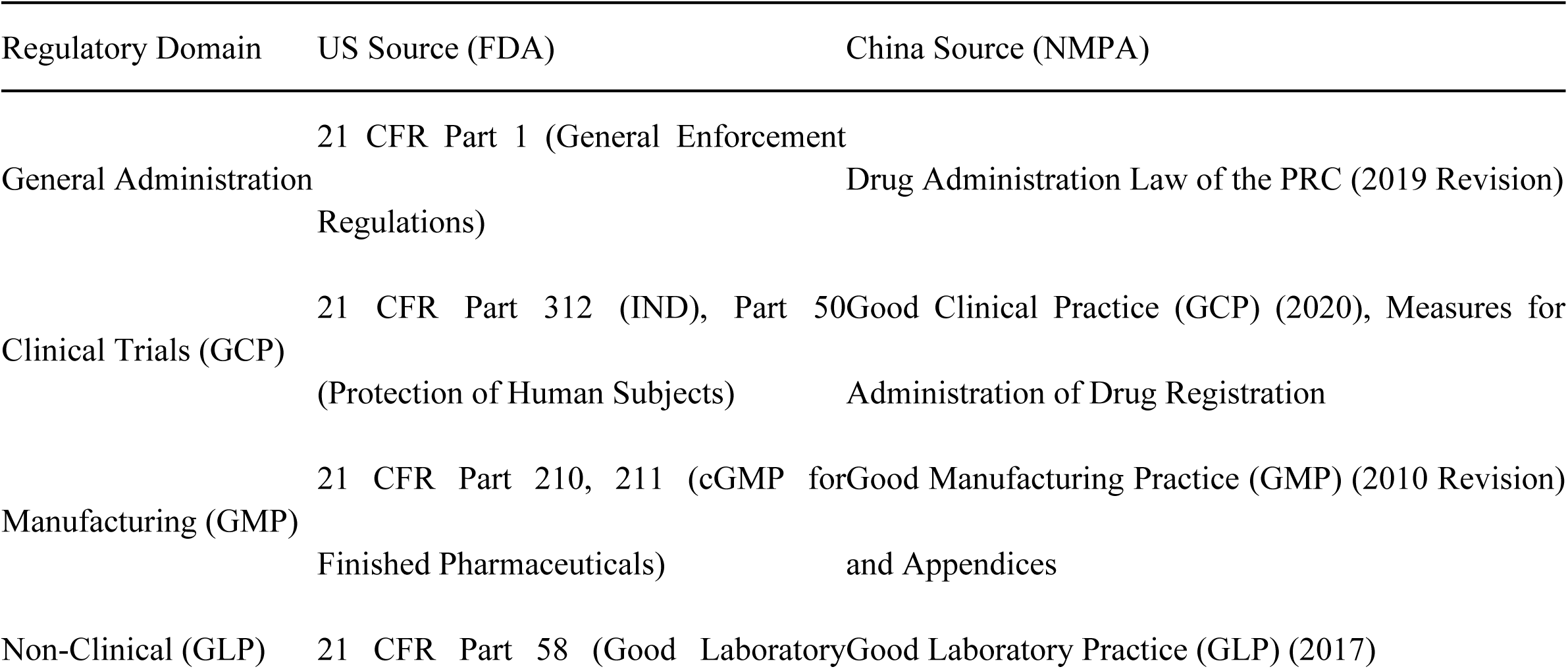
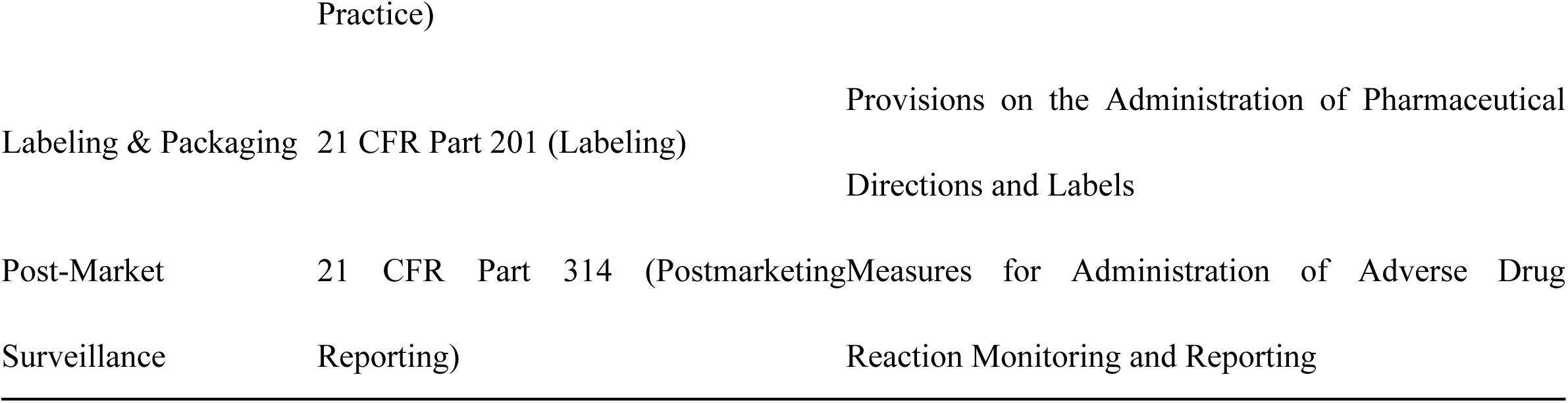
Key regulatory documents and sources included in the Sino-US-DrugQA dataset.

These regulatory documents serve as the exclusive ground-truth source for all question–answer pairs in the dataset. All subsequent dataset construction steps were designed to preserve traceability to the cited regulatory provisions.

### 2.2. Dataset Construction Pipeline

The Sino-US-DrugQA dataset was constructed following a systematic LLM-assisted, human-audited workflow (Fig 1) designed to balance scalability with regulatory accuracy. Automated language models were used to assist in drafting candidate questions, while regulatory interpretation and answer correctness were anchored exclusively in the cited source provisions rather than model inference.

**Fig 1.**
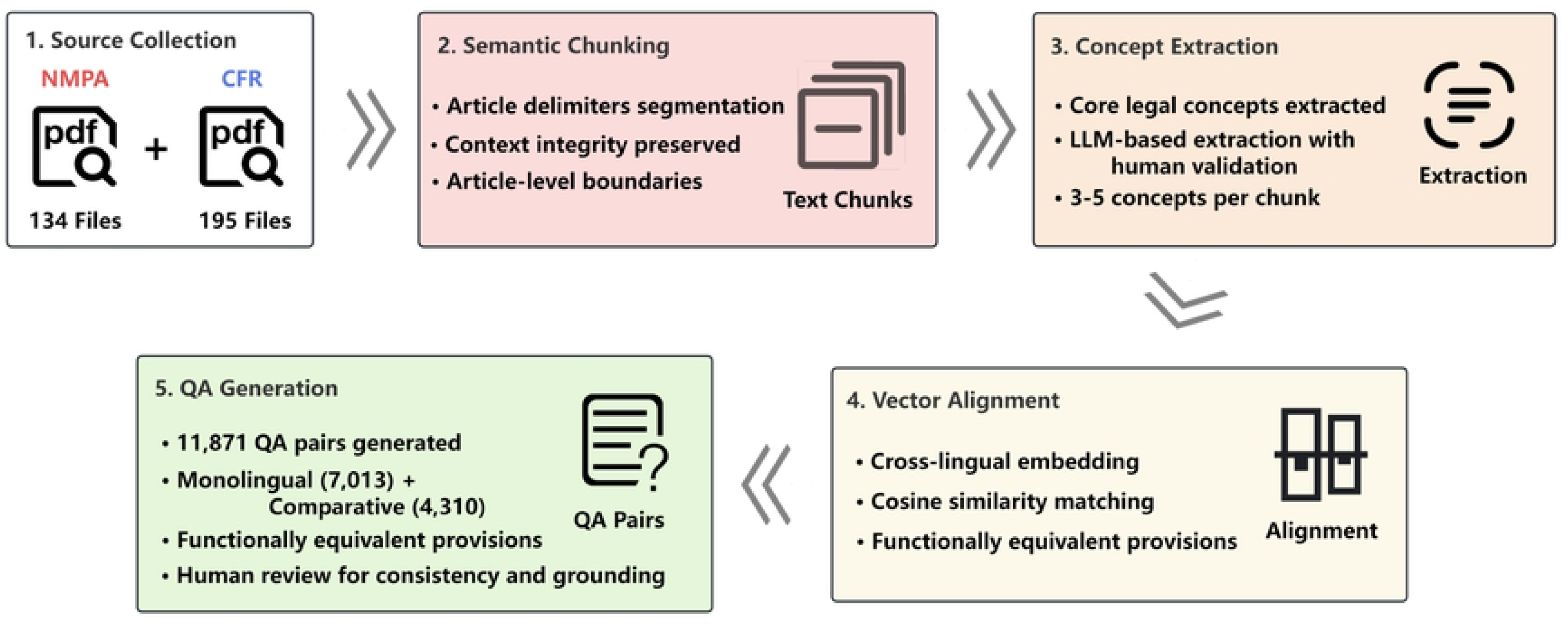
Workflow of the Sino-US-DrugQA dataset construction process, illustrating the five-stage pipeline from source collection to question-answer pair generation.

The dataset construction pipeline consists of five sequential stages:

1. **Source Collection:** Authoritative regulatory texts were collected from official NMPA and eCFR portals, ensuring traceability to publicly accessible, regulator-issued source documents.
2. **Semantic Chunking:** Documents were segmented into provision-level semantic chunks using explicit article delimiters (e.g., “Article 12”, “§ 211.68”) to preserve contextual integrity and avoid cross-article information leakage.
3. **Concept Extraction:** Core regulatory concepts (e.g., adverse event reporting timelines or manufacturing documentation requirements) were identified within each chunk as abstract, compliance-relevant units intended to support comparison, rather than as formal legal interpretations.
4. **Vector Alignment:** To enable cross-jurisdictional comparative reasoning, extracted concepts were mapped using embedding-based similarity methods to identify candidate pairs of functionally corresponding provisions across languages (e.g., aligning US “Lot number” with CN “Batch number”). This alignment step served solely as a retrieval and pairing mechanism and did not determine regulatory equivalence or answer correctness.
5. **QA Generation:** Question–answer pairs were constructed from both aligned and monolingual chunks, covering three task types: (i) Monolingual QA for factual retrieval within a single jurisdiction; (ii) Comparative QA for explicit comparison of regulatory requirements across jurisdictions; and (iii) Parallel QA, in which similar questions were posed independently for each jurisdiction to assess consistency.

Automated language models were used only to assist in drafting question text and candidate answer options. They were not used to determine final regulatory interpretations, alignment decisions, or reference answers. All reference answers were explicitly grounded in the cited regulatory provisions.

### 2.3. Data Quality Assurance and Human Validation

To mitigate risks associated with large-scale automated question–answer generation, we implemented a multi-stage quality assurance protocol that combines automated filtering with targeted expert auditing, rather than exhaustive manual annotation of all items.

**Automated filtering:** All model-generated draft questions were first subjected to automated quality checks. Items with low semantic relevance scores, unclear regulatory grounding, or ambiguous answer choices were excluded prior to expert review.
**Expert spot-check validation:** Given the scale of the dataset, we adopted a random spot-check auditing strategy for human validation. A subset of 500 question–answer pairs was randomly sampled from the full dataset. Two independent regulatory affairs experts, each with more than five years of professional experience in pharmaceutical regulation, reviewed the sampled items for factual correctness, regulatory relevance, and logical consistency with respect to the cited source provisions.
**Inter-annotator agreement:** Agreement between model-generated answers and expert-reviewed corrections was quantified using Cohen’s Kappa coefficient. The resulting κ value of 0.85 indicates strong agreement within the validated subset in expert judgments within the audited subset [17], suggesting that the automated generation pipeline produces regulatory questions of generally high quality.

We emphasize that this expert review was conducted as a quality auditing procedure rather than exhaustive verification of all dataset items. Accordingly, Sino-US-DrugQA is intended as a benchmarking and evaluation resource rather than a substitute for expert regulatory judgment. Any downstream use - particularly in cross-jurisdictional regulatory decision-making - should remain subject to expert interpretation and oversight.

### 2.4. Dataset Characteristics

The final Sino-US-DrugQA dataset comprises 11,871 multiple-choice question–answer pairs constructed from authoritative regulatory documents issued by the US FDA and China’s NMPA. Table 3 summarizes the core characteristics of the dataset, including task types, language distribution, and regulatory domain coverage.

**Table 3:**
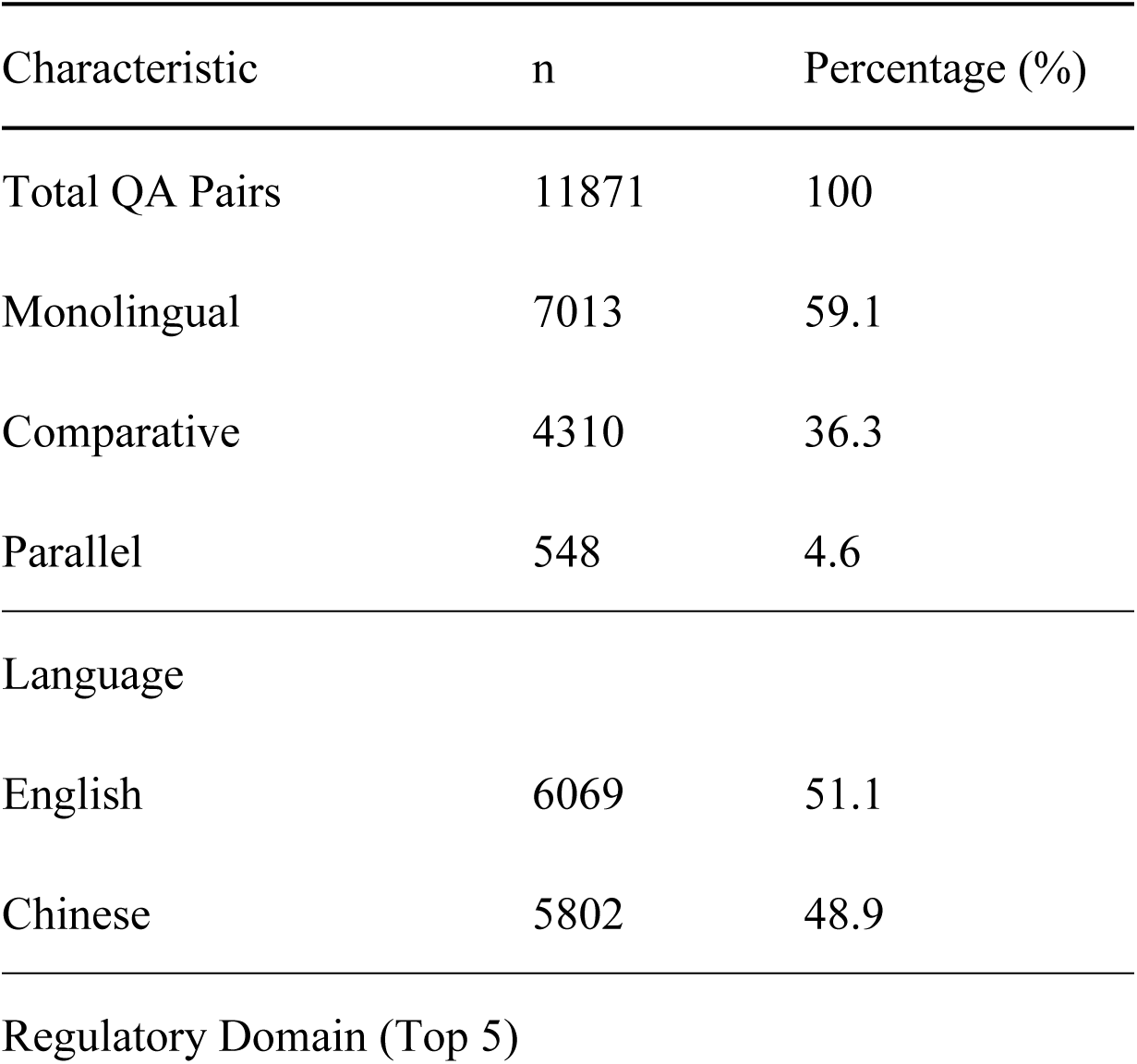
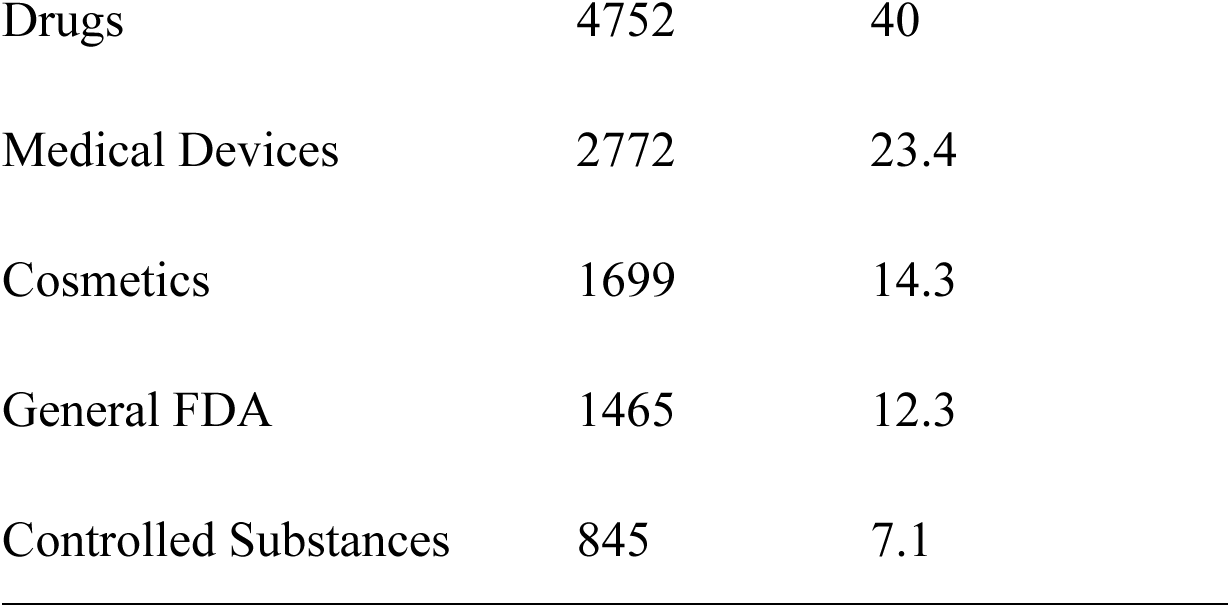
Baseline characteristics of the Sino-US-DrugQA dataset.

| Characteristic | n | Percentage (%) |
| --- | --- | --- |
| Total QA Pairs | 11871 | 100 |
| Monolingual | 7013 | 59.1 |
| Comparative | 4310 | 36.3 |
| Parallel | 548 | 4.6 |
| Language |  |  |
| English | 6069 | 51.1 |
| Chinese | 5802 | 48.9 |
| Regulatory Domain (Top 5) |  |  |
| Drugs | 4752 | 40 |
| Medical Devices | 2772 | 23.4 |
| Cosmetics | 1699 | 14.3 |
| General FDA | 1465 | 12.3 |
| Controlled Substances | 845 | 7.1 |

From a task perspective, Monolingual QA accounts for the majority of the dataset (59.1%), reflecting common real-world use cases in which regulatory professionals query requirements within a single jurisdiction. Comparative QA (36.3%) explicitly targets cross-jurisdictional reasoning by requiring models to compare regulatory requirements across the US and China, while Parallel QA (4.6%) probes consistency by posing structurally similar questions independently in each jurisdiction. Although Parallel QA represents a smaller proportion of the dataset, it is intentionally designed as a diagnostic task to assess cross-lingual consistency rather than to contribute substantially to overall accuracy.

The dataset is linguistically balanced, with 51.1% of questions in English and 48.9% in Chinese, enabling systematic evaluation of bilingual comprehension and cross-lingual consistency in regulatory reasoning, particularly in scenarios involving jurisdiction-specific terminology and administrative structures.

#### 2.4.1. Regulatory domain taxonomy

To facilitate fine-grained performance analysis across different regulatory contexts, all questions were assigned to one of five high-level regulatory domains based on the primary regulatory object and thematic focus of the underlying source documents. This taxonomy was defined during dataset construction and is intentionally coarse-grained, aiming to support comparative capability analysis rather than detailed legal categorization. Each question is associated with a single primary domain.

The five regulatory domains are defined as follows:

**Drugs:** Regulation across the full pharmaceutical product lifecycle, including drug registration and approval pathways, clinical trials, good manufacturing practice (GMP), pharmacovigilance and adverse event reporting, labeling and advertising compliance, post-approval changes, and post-market surveillance.
**Medical Devices:** Regulation of medical devices and in vitro diagnostics, covering device classification, registration or filing requirements, quality management systems (e.g., QMS/QSR/ISO 13485), labeling and UDI, post-market adverse event reporting, recalls and corrective actions, clinical evaluation, and manufacturing or import compliance.
**Cosmetics:** Regulation of cosmetic products and ingredients, including product classification, ingredient compliance and restricted substances, efficacy claims and labeling, safety testing requirements, manufacturing and distribution practices, post-market surveillance, and, where applicable, import or cross-border e-commerce regulation.
**General FDA:** Cross-cutting administrative and institutional rules issued by the FDA that are not limited to a specific product category. This domain includes regulatory framework definitions, inspection and enforcement mechanisms, recordkeeping and documentation requirements, general procedural rules, compliance and penalty frameworks, and mechanisms for regulatory communication or appeal. Functionally corresponding administrative rules from the Chinese regulatory system are included in this category for comparative analysis.
**Controlled Substances:** Regulation of controlled substances, narcotics, psychotropic drugs, and related chemicals, including licensing and quota systems, storage and security requirements, prescription and distribution controls, transportation and inventory management, reporting obligations, and penalties for non-compliance, as well as requirements relevant to manufacturing or research use.

As illustrated in Fig 2, the distribution of regulatory domains reflects practical regulatory workloads in cross-border pharmaceutical activities. Drug-related regulations constitute the largest proportion of the dataset (40.0%), followed by Medical Devices (23.4%) and Cosmetics (14.3%), while General FDA (12.3%) and Controlled_Substances (7.1%) represent more specialized but compliance-critical regulatory areas.

**Fig 2.**
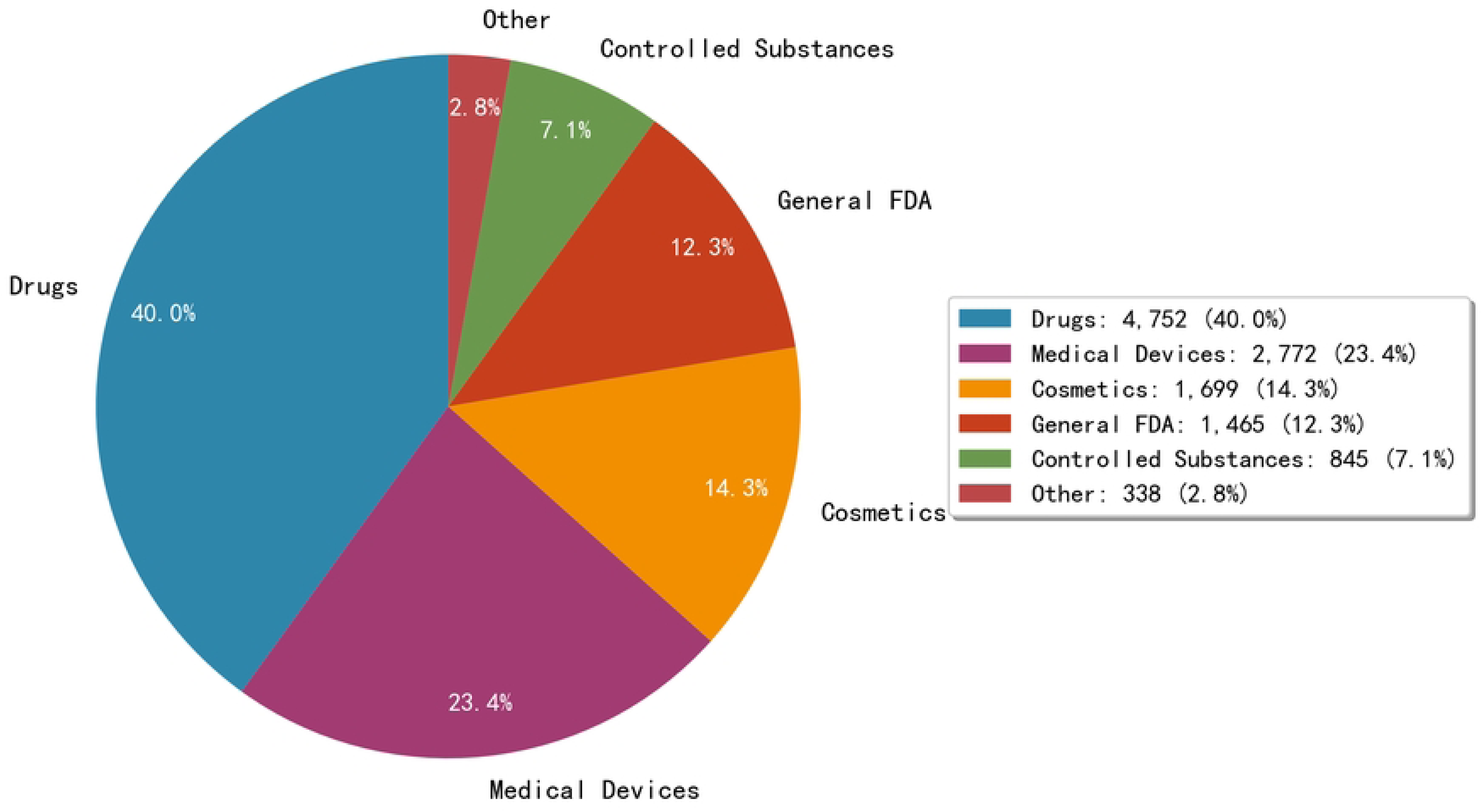
Distribution of regulatory domains within the dataset.

The predominance of the Drugs domain is primarily attributable to the breadth and regulatory density of pharmaceutical regulation across the full product lifecycle, encompassing clinical trials, manufacturing, pharmacovigilance, labeling, post-approval changes, and post-market surveillance. In addition, drug-related regulations exhibit a higher frequency of jurisdiction-specific procedural divergence between the US and China, making them a central focus of cross-jurisdictional compliance analysis in real-world regulatory practice.

This domain distribution enables systematic analysis of whether LLM performance varies across regulatory contexts with differing degrees of international harmonization and jurisdiction-specific administrative complexity.

#### 2.5. Models Evaluated

We selected four representative large language models to establish a diverse and practically relevant evaluation baseline. The selection was guided by three considerations: (i) coverage of both proprietary and open-weight models, (ii) demonstrated capability in English–Chinese bilingual tasks, and (iii) diversity in model design focus, including general-purpose and reasoning-oriented architectures. The evaluated models include:

**GPT-5.2 (OpenAI)** A proprietary large language model with strong general reasoning and instruction-following capabilities, commonly used as a reference point in regulatory and medical NLP benchmarks.
**Gemini-3-flash (Google DeepMind):** A proprietary model optimized for efficiency and long-context understanding, representing recent advances in multimodal and high-throughput inference systems.
**Qwen-3-235B (Alibaba Cloud):** An open-weight large-scale model with documented strength in English–Chinese bilingual understanding, making it particularly relevant for cross-lingual regulatory analysis.
**DeepSeek-V3.2 (DeepSeek AI):** An open-weight model designed with an emphasis on reasoning and technical domains, representing models optimized for structured and analytical tasks.

Together, these models provide a balanced evaluation set spanning different deployment paradigms, linguistic capabilities, and reasoning styles, without targeting exhaustive coverage of all available LLMs.

#### 2.6. Evaluation Protocol

To facilitate reproducibility and community adoption, we developed a standardized evaluation harness that adopts design principles commonly used in recent large-scale LLM benchmarks, including MMLU [18]. Specifically, our evaluation framework follows MMLU-style practices in terms of unified prompt templates, strict output format enforcement (JSON-based parsing), and consistent zero-shot and few-shot evaluation protocols across models. The evaluation code and dataset splits (dev/test) are hosted on GitHub.

Before the main analysis, a pilot evaluation using 10 randomly selected questions was conducted. This pilot phase was used solely for prompt formatting validation rather than performance estimation. And it aimed to refine the instruction prompts and verify alignment between AI outputs and expected JSON formats. Adjustments were made to the system prompts to ensure that the models consistently generated context-appropriate recommendations and valid JSON structures.

##### 2.6.1. Prompt Engineering

We employed a standardized, instruction-based prompting strategy designed to minimize output variability and ensure reproducible evaluation across models. The complete prompt templates, including system instructions, user input structure, and strict JSON output schema, are provided in S1 Appendix to support exact replication of the evaluation protocol. All models were evaluated under two settings: Zero-shot, in which only the target question and multiple-choice options were provided, and Five-shot, in which five randomly sampled exemplars from the development set were prepended to the input.

In both settings, models were instructed to assume the role of a regulatory expert familiar with US FDA and Chinese NMPA regulations, analyze the regulatory context, and select the most appropriate answer from the given options. To facilitate automated parsing and downstream evaluation, responses were constrained to a strict JSON schema specifying the selected option and a brief textual justification. The complete prompt templates and JSON output schema are provided in S1 Appendix.

For all evaluations, the temperature parameter was fixed at 0 to minimize stochastic variation and ensure determinism. The generated reasoning field was not used for accuracy scoring and was collected solely for qualitative error analysis.

#### 2.7. Statistical Analysis

Statistical analyses were performed using IBM SPSS Statistics version 25.0 and Python (scipy and statsmodels libraries). Descriptive statistics were presented as counts and percentages for categorical variables. The primary outcome measure was Accuracy. Statistical significance of performance differences between models was assessed using McNemar’s test for paired nominal data. All McNemar’s tests were conducted on paired model predictions across the full evaluation set (n = 11,871), without any manual intervention.These statistical tests were used exclusively to assess performance differences between models and are independent of the qualitative error analyses reported later. Agreement beyond chance between model predictions and ground-truth labels was additionally reported using Cohen’s Kappa as a complementary metric.

#### 2.8. Data Availability and Reproducibility

The complete Sino-US-DrugQA dataset is publicly available to the research community. The dataset, along with the standardized evaluation harness, prompt templates, and baseline scoring scripts used in this study, can be accessed via our GitHub repository at https://github.com/DodgeLU/Sino-US-DrugQA. Detailed prompt specifications and output schemas required for exact replication of the evaluation protocol are also provided in S1 Appendix.

### 3. Results

#### 3.1. Overall Performance

A total of 11,871 question-answer pairs were analyzed. Overall accuracy reflects a task-distribution–weighted average across Monolingual, Comparative, and Parallel QA tasks, reflecting the task distribution of the benchmark, in which monolingual regulatory queries constitute the majority of instances. The overall concordance rate between model-generated recommendations and ground truth answers is summarized in Table 4.

**Table 4:** Overall accuracy and performance metrics for all evaluated models (Zero-shot Setting).

|  | DeepSeek-V3.2 | GPT-5.2 | Qwen-3-235B | Gemini-3-flash |
| --- | --- | --- | --- | --- |
| CN (%) | 83.95% | 82.30% | 84.83% | 87.62% |
| EN (%) | 77.26% | 75.80% | 75.45% | 81.53% |
| Comparative (%) | 76.80% | 73.83% | 75.36% | 79.63% |
| Monolingual (%) | 82.86% | 82.65% | 83.12% | 87.61% |
| Parallel (%) | 79.93% | 72.45% | 77.37% | 83.21% |
| Overall Accuracy (%) | 80.53% | 78.97% | 80.04% | 84.51% |
| $\kappa$ (95% CI) | 0.699 (0.688–0.710) | 0.687 (0.677–0.697) | 0.699 (0.689–0.711) | 0.764 (0.754–0.774) |

Gemini-3-flash demonstrated the highest overall accuracy (84.51%), outperforming the other evaluated models across all task categories. DeepSeek-V3.2 (80.53%) and Qwen-3-235B (80.04%) also exhibited strong performance, while GPT-5.2 achieved an overall accuracy of 78.97%.

In addition to accuracy, agreement between model-generated answers and reference answers grounded in the cited regulatory provisions was evaluated using Cohen’s Kappa. As shown in Table 4, Gemini-3-flash achieved the highest κ value (0.764, 95% CI: 0.754–0.774), while the other models exhibited κ values in the range of 0.687–0.699.

These κ values indicate substantial agreement under the benchmark’s reference-answer framework, suggesting that performance differences across models are not solely driven by class imbalance or majority-class effects. We note that κ reflects consistency with the constructed benchmark rather than independent validation of regulatory correctness, and should therefore be interpreted as a complementary metric to accuracy rather than a definitive measure of regulatory validity.

#### 3.2. Performance by Task Type

Subgroup analysis by task type revealed significant variability in model performance (Fig 3). All models scored higher on Monolingual QA tasks compared to Comparative QA tasks:

1. Gemini-3-flash: Monolingual 87.61% vs. Comparative 79.63% (Difference: ∼8.0%)
2. DeepSeek-V3.2: Monolingual 82.86% vs. Comparative 76.80% (Difference: ∼6.1%)
3. Qwen-3-235B: Monolingual 83.12% vs. Comparative 75.36% (Difference: ∼7.8%)
4. GPT-5.2: Monolingual 82.65% vs. Comparative 73.83% (Difference: ∼8.8%)

**Fig 3.**
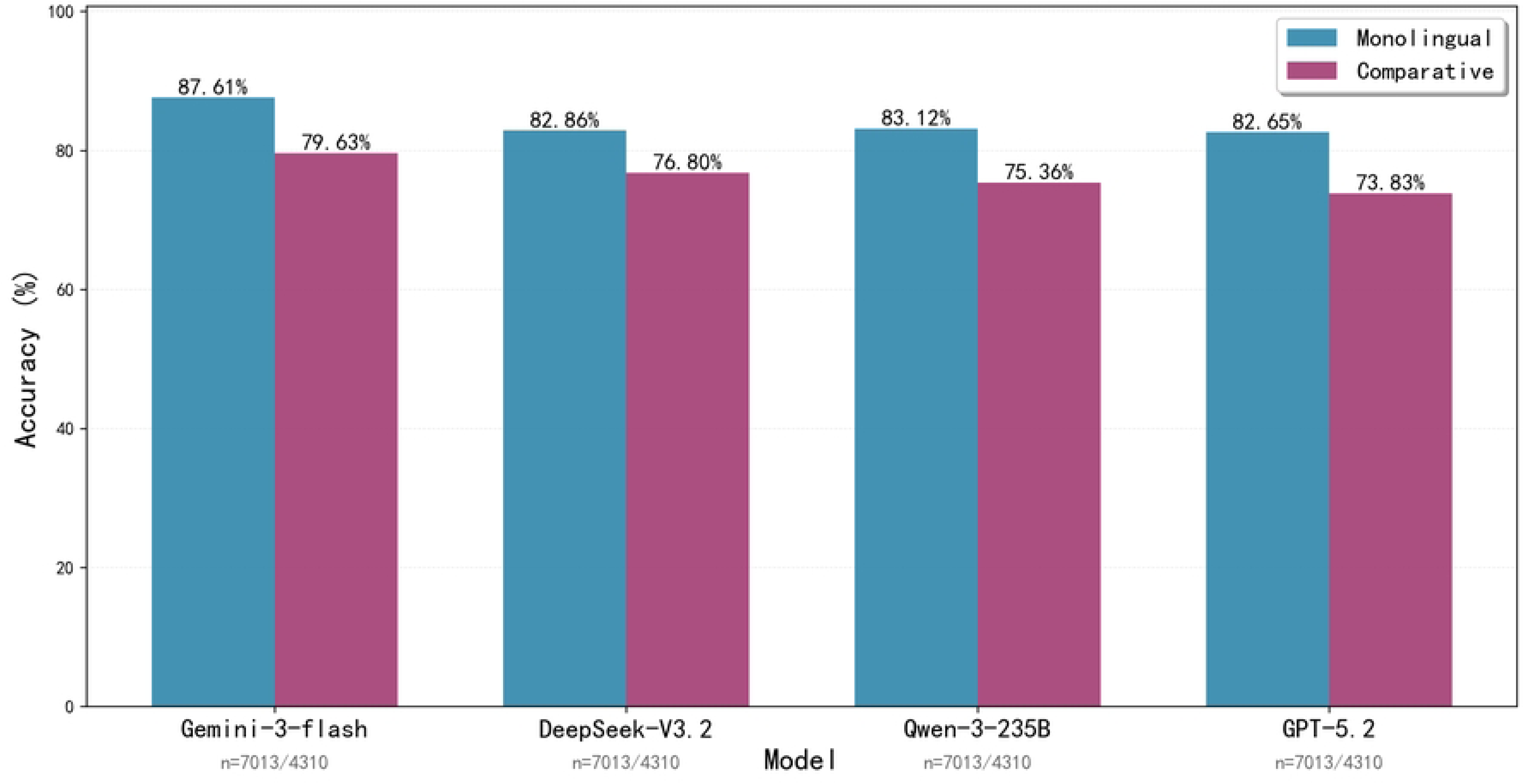
Accuracy comparison by task type (Monolingual vs. Comparative).

This performance degradation in comparative tasks - which we term the cross-jurisdictional performance gap - indicates that while current LLMs are proficient at retrieving and understanding isolated regulatory provisions within a single legal system, they face challenges when simultaneously holding two distinct legal frameworks in context and performing logical deduction to identify differences.

#### 3.3. Impact of In-Context Learning

Building upon the Zero-shot baseline results reported above, we further evaluated the impact of in-context learning on regulatory reasoning. We compared model performance under Zero-shot and Five-shot settings. As illustrated in Fig 4, providing five relevant examples generally improved performance across all models, particularly in the Comparative QA category.

**Fig 4.**
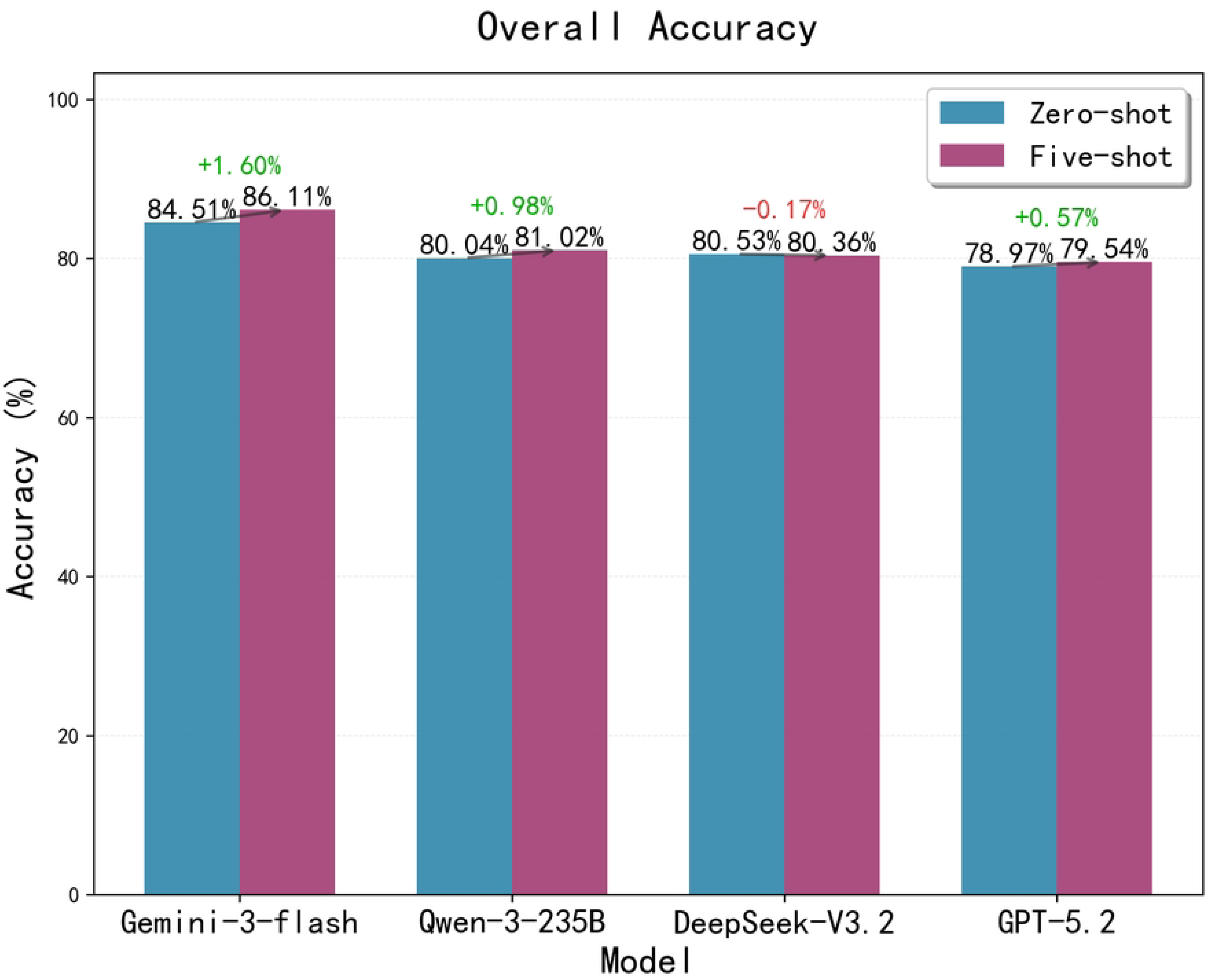
(a) Overall accuracy comparison between Zero-shot and Five-shot settings across evaluated models; (b) Comparative QA accuracy comparison between Zero-shot and Five-shot settings across evaluated models.

However, even with few-shot prompting, the cross-jurisdictional performance gap persisted. This suggests that while examples help models understand the task format and output structure, they do not fully bridge the fundamental knowledge gap required for complex cross-border alignment. Notably, DeepSeek-V3.2 exhibited a slight performance degradation under the Five-shot setting, particularly for Comparative QA, indicating potential sensitivity to exemplar selection or prompt overfitting. This observation suggests that in-context learning does not uniformly benefit all model architectures in cross-jurisdictional regulatory reasoning.

#### 3.4. Performance Across Regulatory Functional Sub-domains

To further understand where models succeed or fail within regulatory reasoning tasks, we conducted a fine-grained analysis across regulatory functional sub-domains. These sub-domains represent recurring regulatory activities that cut across product categories, such as registration procedures, labeling requirements, and post-market obligations, rather than the regulatory object itself.

The highest accuracy was consistently observed in GMP and GLP-related tasks. These domains benefit from a high degree of international harmonization through frameworks such as ICH Q7 and OECD GLP principles [19,20]. They emphasize technical standards and procedural consistency, which appear to align well with the training distributions of current LLMs.

In contrast, performance was lowest in Labeling and General Administrative procedures. These functional areas are characterized by jurisdiction-specific formatting rules, documentation practices, and administrative thresholds (e.g., mandatory Chinese labeling elements versus FDA “Drug Facts” panels). Such requirements often lack direct cross-jurisdictional counterparts, increasing the difficulty of alignment and comparison.

Importantly, this analysis focuses on functional regulatory tasks rather than product-based regulatory categories. As such, performance trends observed here are not intended to correspond directly to the high-level regulatory domains reported in Section 3.5.

#### 3.5. Performance by Regulatory Object Domain

Complementing the functional analysis above, we next examined model performance across the five high-level regulatory domains defined in Section 2.4, which group questions according to the primary regulatory object (e.g., drugs, medical devices, controlled substances).

As illustrated in Fig 5, the highest overall accuracy was observed in the Drugs and Medical Devices domains. These areas are relatively well represented in public regulatory guidance and international standards (e.g., ICH, ISO 13485), which may contribute to more robust model representations.

**Fig 5.**
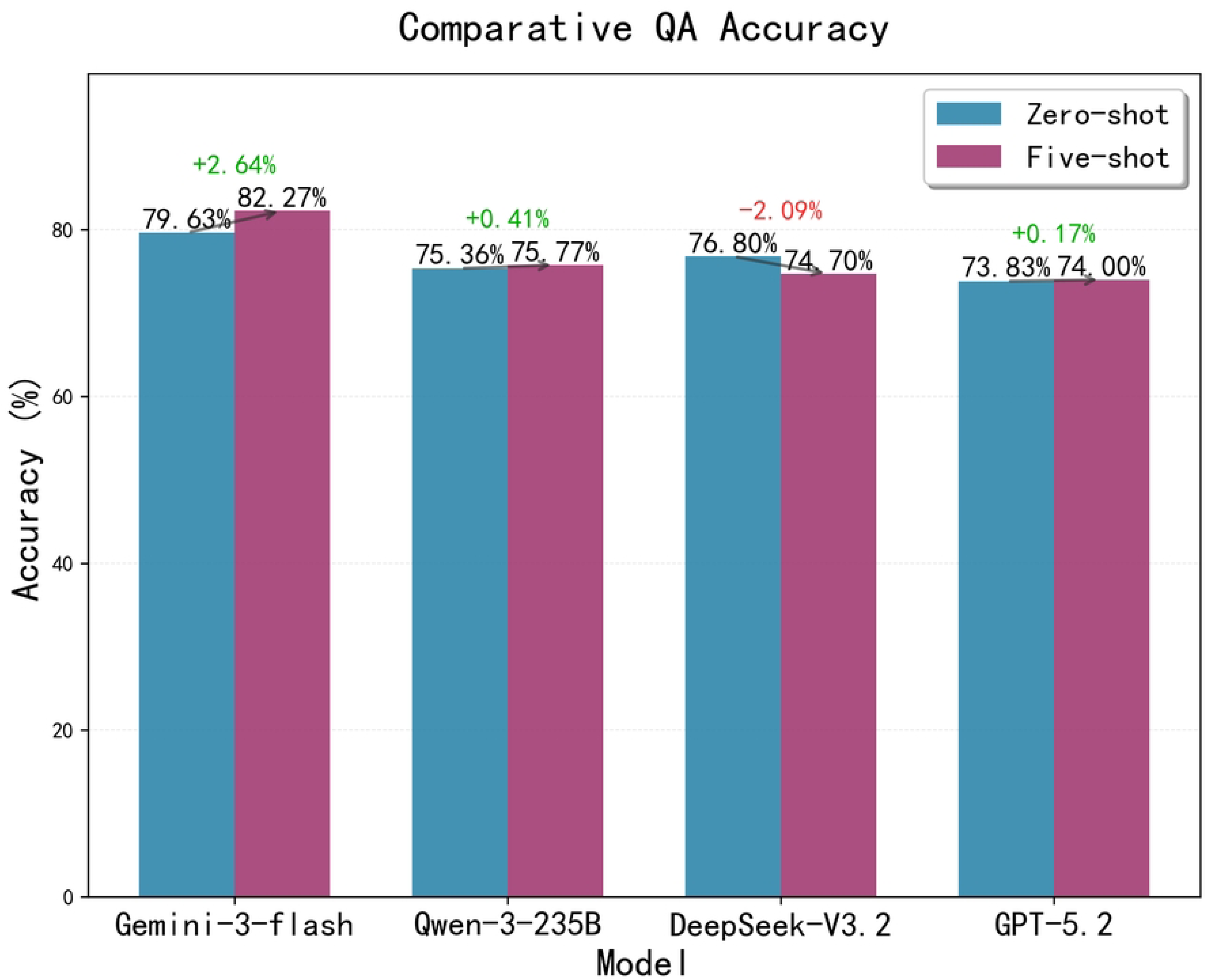
Heatmap of model accuracy across different regulatory domains.

By contrast, performance was lowest in the Controlled Substances and General FDA domains. Unlike labeling or registration as functional tasks, these domains are dominated by jurisdiction-specific administrative procedures, enforcement mechanisms, and penal provisions. Regulatory concepts in these areas are often tightly coupled to national legal frameworks and institutional practices, limiting the transferability of knowledge across jurisdictions.

Notably, lower performance in Controlled Substances does not contradict the findings in Section 3.4. While labeling emerged as a challenging functional task, Controlled Substances represents a regulatory object domain with high legal specificity and limited international harmonization. These two analyses capture different dimensions of regulatory complexity and jointly highlight where cross-jurisdictional alignment is most fragile.

#### 3.6. Statistical Significance and Error Analysis

**Table 5:** McNemar’s Test Significance Matrix.

| Model | DeepSeek-V3.2 | GPT-5.2 | Qwen-3-235B | Gemini-3-flash |
| --- | --- | --- | --- | --- |
| DeepSeek-V3.2 | / | <0.001 | 0.062 | <0.001 |
| GPT-5.2 | <0.001 | / | 0.049 | <0.001 |
| Qwen-3-235B | 0.062 | 0.049 | / | <0.001 |
| Gemini-3-flash | <0.001 | <0.001 | <0.001 | / |

McNemar’s test was applied to paired model predictions on identical question sets to assess whether observed performance differences between models were statistically significant. As a paired nominal test, this analysis evaluates directional differences in error patterns but does not assume independence between models or imply causal superiority.

To better understand common failure modes, we conducted an exploratory qualitative error analysis on a randomly sampled subset of model failures. This analysis was designed as a descriptive auditing procedure rather than a basis for statistical generalization to the full dataset.

A qualitative error analysis of 400 randomly sampled failures in Comparative QA revealed three primary error types:

1. Hallucination (18%): Fabricating regulatory clauses or article numbers.
2. Misalignment (25.5%): Correctly citing the US regulation but failing to retrieve the corresponding Chinese provision (or vice versa).
3. Logical Failure (56.5%): Correct retrieval of relevant regulatory provisions but incorrect comparative deduction, such as reversing which jurisdiction specifies more detailed personnel qualifications, or misjudging the validity of absolute procedural statements across regulatory scopes.

These error categories were defined prior to analysis and applied consistently across all sampled instances.The prevalence of Misalignment errors underscores the challenge of cross-lingual semantic retrieval and functional concept alignment in the absence of explicit alignment aids.

#### 3.7. Representative Error Examples

To qualitatively illustrate the error patterns identified in Section 3.6, we present two representative error examples directly drawn from the model evaluation outputs and the released Sino-US-DrugQA question set, each traceable via a shared question identifier (id). These examples are intended for explanatory and diagnostic purposes only and are not used to support statistical inference or comparative performance claims.

##### Case 1: Comparative Reasoning Reversal under Overgeneralized Regulatory Interpretation

**Question ID:** CO_P_00001_004

**Question (summary):** Which jurisdiction specifies more detailed personnel qualification requirements for blood product manufacturing?

**Ground Truth:** B - Chinese regulations specify more explicit educational and experience requirements for designated roles, whereas US regulations emphasize general competency principles.

**Model Error (GPT-5.2):** The model selected A, incorrectly attributing greater prescriptive detail to US regulations.

**Analysis:** The model demonstrated correct retrieval of relevant US regulatory provisions (e.g., 21 CFR Parts 600–680) but overgeneralized their practical specificity, leading to a reversal in comparative judgment. This error does not stem from fabricated citations, but rather from comparison-specific deductive failure, where qualitative regulatory language (“appropriate education, training, and experience”) was implicitly equated with explicit degree- and tenure-based requirements present in the Chinese framework. Such errors were frequently observed in comparative questions requiring assessment of relative regulatory granularity rather than absolute presence of requirements.

##### Case 2: Scope Sensitivity Failure Triggered by Absolute Language in Comparative Statements

**Question ID:** CO_P_00051_002

**Question (summary):** Two statements regarding externally supplied herbal extracts and written procedures for identity and contamination control; determine which combination is correct.

**Ground Truth:** A—both statements are correct under their respective regulatory systems.

**Model Error (GPT-5.2):** The model selected B, judging the second statement incorrect due to the use of the word “always.“

**Analysis:** The model correctly identified the regulatory logic underlying both Chinese and US systems but rejected the second statement based on an over-sensitive interpretation of absolute wording (“always”). In doing so, it failed to account for the fact that the statement accurately reflects binding procedural requirements within the applicable regulatory scope (e.g., written SOPs developed by qualified personnel under US cGMP). This represents a scope and boundary misinterpretation rather than a factual retrieval error, highlighting a common failure mode in comparative true/false questions where models apply overly strict linguistic heuristics instead of regulatory-contextual reasoning.

### 4. Discussion

#### 4.1. Cross-jurisdictional performance gap

This study introduces Sino-US-DrugQA to address the lack of specialized benchmarks for cross-jurisdictional pharmaceutical regulation. Across four evaluated LLMs, a consistent pattern was observed: while models achieved high accuracy on monolingual retrieval tasks, performance decreased by approximately 6–9 percentage points when tasked with comparative reasoning.

This cross-jurisdictional performance gap reflects a key limitation of current LLMs in regulatory settings. Compared with general-domain benchmarks (e.g., MMLU) or monolingual legal benchmarks (e.g., LexGLUE), cross-jurisdictional compliance requires a model to (1) retrieve provisions from two distinct regulatory systems, (2) align them despite linguistic and structural differences, and (3) perform comparison-specific deduction (e.g., “which is stricter/longer/shorter”). The observed drop suggests that alignment and comparison, rather than retrieval alone, are the primary bottlenecks.

#### 4.2. Cross-lingual Translation Alone Is Insufficient for Cross-jurisdictional Regulatory Alignment

A common assumption in multilingual regulatory question answering is that accurate cross-lingual translation can largely bridge performance gaps across jurisdictions [21]. Our findings suggest that, while translation is a necessary component, it is insufficient on its own to support robust cross-jurisdictional regulatory reasoning.

First, quantitative results indicate that performance gaps persist even when linguistic barriers are minimized. In the Parallel QA setting, where structurally equivalent questions are independently posed in Chinese and English, model accuracy remains substantially lower than in monolingual tasks. This suggests that errors cannot be fully attributed to surface-level translation difficulty.

Second, comparative tasks that explicitly require cross-jurisdictional reasoning exhibit a sharper performance drop than both monolingual and parallel tasks. Notably, this degradation is observed even in domains where terminology is relatively standardized, such as GMP-related questions, indicating that jurisdiction-specific regulatory logic - rather than vocabulary alone - poses a significant challenge.

Third, qualitative error analysis reveals recurring failure patterns that are not resolvable through improved translation. For example, models frequently conflate FDA procedural mechanisms with NMPA administrative processes, misinterpret approval pathways as interchangeable, or incorrectly assume equivalence between regulatory concepts that are superficially similar but legally distinct. These errors persist even when the underlying regulatory texts are accurately translated.

Taken together, these results demonstrate that cross-lingual translation addresses only the linguistic layer of the problem. Effective cross-jurisdictional regulatory alignment additionally requires models to internalize jurisdiction-specific regulatory structures, institutional roles, and compliance logic. Our benchmark highlights this limitation by isolating scenarios in which linguistic equivalence does not imply regulatory equivalence.

#### 4.3. Regulatory hallucination and compliance risk

The error typology indicates that hallucination and misalignment, as observed in the benchmark setting, are potentially consequential beyond purely academic evaluation. In real-world regulatory practice, errors such as fabricated clauses, mis-cited article numbers, or incorrect comparative assumptions regarding regulatory obligations may plausibly contribute to compliance risk if relied upon without verification. Prior work on generative AI in healthcare has emphasized the need for governance, oversight, and risk controls when deploying LLMs in safety-critical environments [8,9]. Our findings are consistent with this broader literature, although the present study does not attempt to quantify downstream regulatory impact.

In cross-border regulatory affairs, where legal obligations differ structurally across jurisdictions, such model failure modes may be particularly problematic when LLM outputs are used as preliminary decision-support inputs. Examples of potential downstream consequences include misinterpretation of jurisdiction-specific requirements, incorrect comparative assumptions, or reliance on superseded regulatory provisions, rather than direct operational errors.

Accordingly, evaluation should be framed in terms of decision support rather than autonomous decision-making. Sino-US-DrugQA is intended to surface model-internal failure patterns systematically by quantifying where models fail (e.g., comparison-specific deduction versus cross-lingual concept alignment), and by providing analytically defined error categories to inform mitigation strategies such as citation checking, retrieval grounding, and expert review, rather than to establish causal links to real-world compliance outcomes.

#### 4.4. From automation to augmentation: a human-in-the-loop workflow

The observed agreement levels (e.g., Cohen’s κ for the top-performing model) suggest that current LLMs can be useful for drafting and first-pass screening in regulatory tasks, particularly for monolingual retrieval. However, the persistent performance gap in comparative tasks supports a conservative deployment posture: LLM outputs in cross-border comparisons should be treated as probabilistic suggestions and must be subject to expert verification.

In practice, a human-in-the-loop workflow is recommended: models can propose candidate differences, cite relevant provisions, and generate draft comparison notes, while regulatory professionals retain responsibility for final interpretation and compliance decisions.

#### 4.5. Positioning Sino-US-DrugQA relative to human performance

A further consideration is how to position this benchmark relative to human expertise. While some studies report strong LLM performance on medical examinations, cross-jurisdictional regulatory comparison imposes additional demands: it requires knowledge of administrative procedures and the ability to reconcile non-equivalent categories across jurisdictions. Therefore, high performance on general medical QA or legal QA does not necessarily translate into safe regulatory comparison.

In this context, Sino-US-DrugQA is not intended to approximate or replace expert-level regulatory judgment, but rather to evaluate whether LLMs can reliably support preliminary regulatory information retrieval and comparison tasks under expert supervision. By explicitly exposing failure modes related to cross-jurisdictional alignment and comparative deduction, the benchmark helps delineate which subtasks may be amenable to cautious automation and which remain dependent on human expertise. Accordingly, Sino-US-DrugQA is positioned as a decision-support–oriented evaluation resource, rather than a proxy for human regulatory performance.

#### 4.6. Limitations and future directions

This study has several limitations. First, although we implemented spot-check expert validation, portions of the dataset construction relied on automated pipelines, which may introduce systematic biases. Specifically, expert review was conducted as a quality auditing mechanism to assess the reliability of the automated generation pipeline, rather than as exhaustive item-level verification. Second, regulatory “ground truth” can be context-dependent; we mitigated this by focusing on questions grounded in explicit regulatory text rather than discretionary interpretation, but edge cases remain. Third, the baseline evaluation currently covers a limited set of models and prompting settings; future work should expand model coverage and report few-shot results to assess the stability of improvements under in-context learning. Finally, regulatory landscapes evolve rapidly; periodic dataset maintenance and versioning will be necessary to keep the benchmark aligned with current regulations.

## 5. Conclusions

This study introduces Sino-US-DrugQA, a bilingual benchmark dataset for evaluating large language models in cross-jurisdictional pharmaceutical regulation. The dataset is paired with an open evaluation harness to support transparent, reproducible benchmarking and to lower the barrier for follow-up research in regulatory science. Across baseline evaluations, models performed substantially better on monolingual regulatory questions than on cross-jurisdictional comparative questions, indicating a consistent performance gap in tasks that require cross-jurisdictional comparison and regulatory concept alignment. In practical terms, these results support a conservative use case: current LLMs are best positioned as drafting and screening assistants, while cross-border compliance decisions should remain subject to expert review.

Sino-US-DrugQA is intended as a community resource. Future work should (1) report few-shot results to quantify the extent to which in-context learning improves comparative reasoning, (2) expand model coverage and evaluate stability under paraphrased queries, and (3) maintain and version the benchmark to reflect evolving regulatory landscapes. We invite researchers and practitioners to use the dataset and evaluation scripts to track progress and to develop safer, more reliable regulatory intelligence tools.

## Data Availability

The dataset and evaluation scripts are available at https://github.com/DodgeLU/Sino-US-DrugQA.

## Acknowledgments

**Conceptualization:** Xuejing Fu, Wentao Lu.

**Data curation:** Wentao Lu.

**Investigation:** Xuejing Fu, Xuejing Fu.

**Methodology:** Xuejing Fu, Zhen Chen, Wentao Lu.

**Resources:** Xuejing Fu, Zhen Chen.

**Software:** Wentao Lu.

**Supervision:** Xuejing Fu.

**Validation:** Xuejing Fu, Zhen Chen.

**Writing – original draft:** Xuejing Fu, Wentao Lu.

**Writing – review & editing:** Xuejing Fu, Zhen Chen, Wentao Lu.

## Supporting information

**S1 Appendix.**
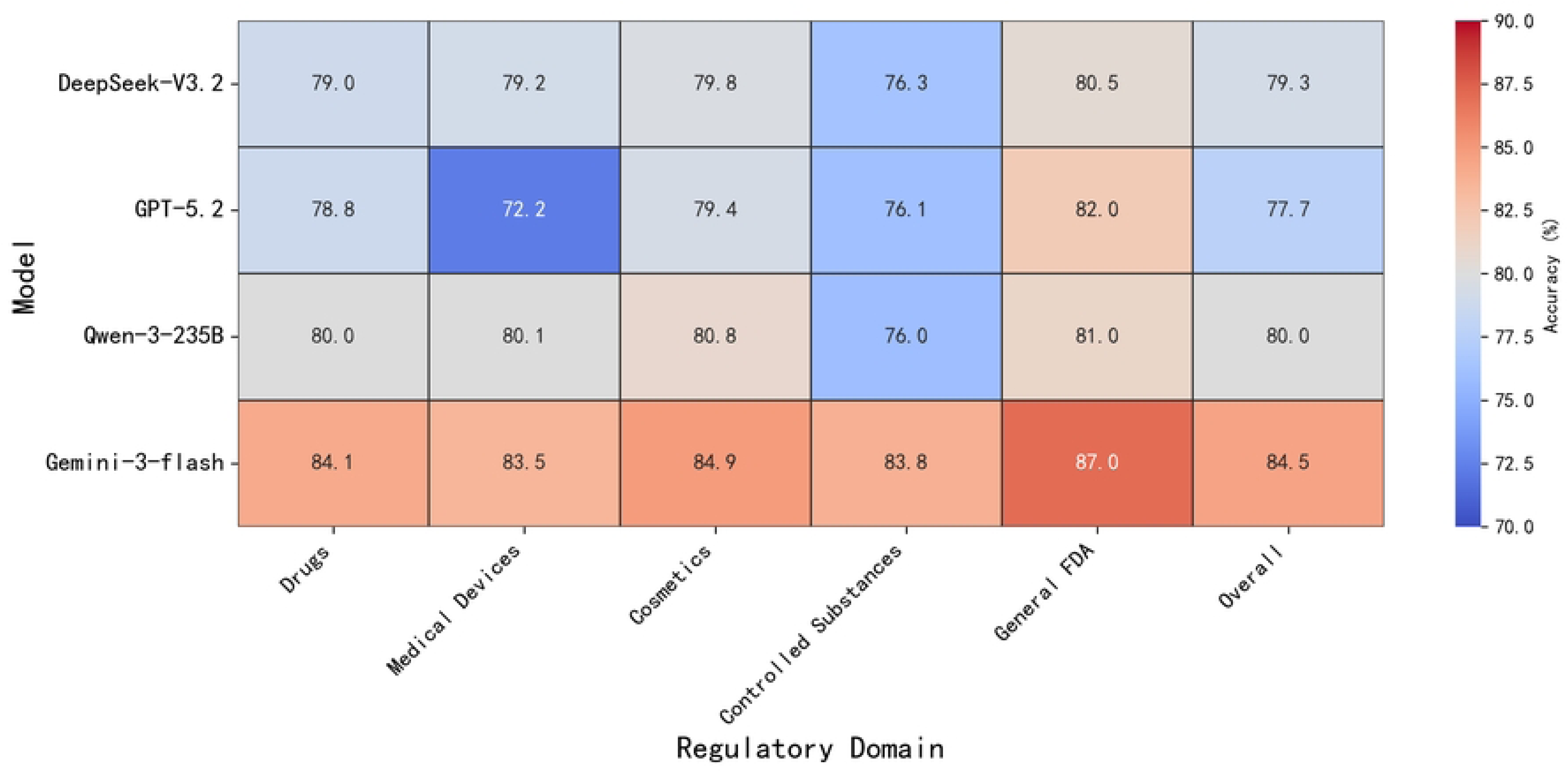
Prompt Templates and Output Schema.

